# Evidence of SARS-CoV-2 transcriptional activity in cardiomyocytes of COVID-19 patients without clinical signs of cardiac involvement

**DOI:** 10.1101/2020.08.24.20170175

**Authors:** Gaetano Pietro Bulfamante, Gianluca Lorenzo Perrucci, Monica Falleni, Elena Sommariva, Delfina Tosi, Carla Martinelli, Paola Songia, Paolo Poggio, Stefano Carugo, Giulio Pompilio

**Affiliations:** Unità di Anatomia Patologica, Dipartimento di Scienze della Salute, Università degli Studi di Milano, Milan, Italy; Struttura Complessa di Anatomia Patologica e Genetica Medica, ASST Santi Paolo e Carlo, Milan, Italy; Unità di Biologia Vascolare e Medicina Rigenerativa, Centro Cardiologico Monzino IRCCS, Milan, Italy; Unità per lo Studio delle Patologie Aortiche, Valvolari e Coronariche, Centro Cardiologico Monzino IRCCS, Milano, Italy; Unità di Cardiologia, Dipartimento di Scienze della Salute, Università degli Studi di Milano, Milan, Italy; Dipartimento di Scienze Cliniche e di Comunità, Università degli Studi di Milano, Milan, Italy

## Abstract

**Background:** Cardiovascular complication in patients affected by novel Coronavirus respiratory disease (COVID-19) are increasingly recognized. However, although a cardiac tropism of SARS-CoV-2 for inflammatory cells in autopsy heart samples of COVID-19 patients has been reported, the presence of the virus in cardiomyocytes has not been documented yet.

**Methods:** We investigated for SARS-CoV-2 presence in heart tissue autopsies of 6 consecutive COVID-19 patients deceased for respiratory failure showing no signs of cardiac involvement and with no history of heart disease. Cardiac autopsy samples were analysed by digital PCR, Western blot, immunohistochemistry, immunofluorescence, RNAScope, and transmission electron microscopy assays.

**Results:** The presence of SARS-CoV-2 into cardiomyocytes was invariably detected. A variable pattern of cardiomyocytes injury was observed, spanning from the absence of cell death and subcellular alterations hallmarks to the intracellular oedema and sarcomere ruptures. In addition, we found active viral transcription in cardiomyocytes, by detecting both sense and antisense SARS-CoV-2 spike RNA.

**Conclusions:** In this analysis of autopsy cases, the presence of SARS-CoV-2 into cardiomyocytes, determining variable patterns of intracellular involvement, has been documented. All these findings suggest the need of a cardiologic surveillance even in survived COVID-19 patients not displaying a cardiac phenotype, in order to monitor potential long-term cardiac sequelae.

## Introduction

Mortality due to the infection of novel severe acute respiratory syndrome coronavirus (SARS-CoV-2) disease (COVID-19)^1^ is still increasing worldwide. Beside SARS-CoV-2 virus lung tropism causing an atypical severe acute respiratory distress syndrome (ARDS),^2^ evidence is now available about the involvement in COVID-19 patients of several other organs, including the cardiovascular system. About 20-40% of hospitalized patients manifest a wide spectrum of symptoms related to heart injury, ranging from mild chest discomfort, palpitations, arrhythmias to cardiogenic shock and fulminant heart failure.^3^ Furthermore, myocardial injury has been documented in more than 50% of deceased patients^4^ with up to 7% of COVID-19 related deaths linked to myocarditis.^5^ To date, it is not clearly understood whether the cause of the cardiovascular involvement of COVID-19 patients is due to SARS-CoV-2 direct cell damage or is rather secondary to the excessive immune system reaction. The systemic hyper-inflammatory status may act as a trigger for an aberrant host immune response mediated by natural killer (NK) cells, macrophages, and T-lymphocytes.^6^ Inflammation-mediated leucocyte adhesion molecules on endothelium may cause coronary dysfunction with ischemia^7^ or acute coronary syndrome.^8^ Alternatively, ARDS-induced hypoxia associated with systemic sepsis, fever, and hypotension may lead to myocardial injury^7,9^ due to the oxygen supply and demand imbalance. On the other hand, SARS-CoV-2 has been found in vascular endothelial cells of patients affected by endotheliitis^10^ as well as in cardiac macrophages of COVID-19 patients with cardiogenic shock and myocarditis.^11,12^ Moreover, viral RNA has been previously reported in COVID-19 patients’ whole heart tissue,^13,14^ regardless of a cardiac phenotype.^15,16^ The cell invasion mechanism of SARS-CoV-2 is mainly mediated by ACE2 receptor^17^, which is highly expressed by several cardiac cell types, such as pericytes,^18^ cardiomyocytes,^19^ and endothelial cells^10,20^. Nonetheless, immunohistochemistry for viral antigen detection in cardiomyocytes has never been shown.^21,22^

Here, we present, for the first time, data on SARS-CoV-2 sense and antisense RNA as well as viral protein presence in cardiomyocytes of autopsy heart samples of 6 patients deceased for COVID-19- dependent severe respiratory insufficiency.

## Materials and Methods

### Data availability

The raw data are available in Zenodo, at https://dx.doi.org/10.5281/zenodo.3974891.

### Patients’ autopsy samples

Autopsies of 6 consecutive non selected patients involved in this study, 5 males and 1 female, aged 54-69 years (average age: 59.5 ± 5.9 years) with a diagnosis of COVID-19, confirmed by CT analysis (**Suppl. Fig. 1**) and polymerase chain reaction (PCR), were performed in a BSL3 equipped autopsy room at the Unit of Pathology of “ASST Santi Paolo e Carlo” of Milan. Examiners worn clad in all necessary DPIs. Examiners followed the recently published guidelines for performing autopsies and samples with suspected COVID-19 (SIAPEC-IAP).^23^ The autopsy examination was performed as soon as possible, by following the recently published recommendations. The period from death to post-mortem autopsy ranged from 1 to 2 hours, documented instrumentally with a continuous and flat ECG for 20 minutes.

All patients had no clinical signs of heart damage and died for SARS-CoV-2-dependent respiratory failure after hospitalization in the Intensive Care Unit. In **Table 1** clinical characteristics of all 6 patients involved in this study have been summarized. The autopsy included complete histopathology and virology evaluation. Clinical records were checked for clinical conditions at hospitalization.

**Table 1.** Clinical data of the analysed patients.

| **Patient** | **1** | **2** | **3** | **4** | **5** | **6** |
| --- | --- | --- | --- | --- | --- | --- |
| **Average age (years)** | 59.5 ± 5.9 |  |  |  |  |  |
| **Sex** | M | M | M | M | M | F |
| **Cause of death** | Respiratory failure | Respiratory failure | Respiratory failure | Respiratory failure | Respiratory failure | Respiratory failure |
| **Previously myocardial damages** | // | // | // | // | // | Mildly hypertrophic LV. EF>50%. |
| **RCP (mg/L)** | 134.4 | 67.7 | 118.1 | 53.8 | 82.8 | 135 |
| **D-DIMER (ng/mL)** | 2242 | 507 | 783 | 583 | 2987 | 906 |
| **TROPONIN I ultrasensitive (ng/mL)** | <0.012 | 0.022 | <0.012 | <0.012 | <0.012 | <0.012 |
| **Hypertension** | YES | NO | NO | NO | NO | YES |
| **Hypercholesterolemia** | YES | NO | NO | YES | NO | NO |
| **Diabetes** | NO | NO | NO | NO | NO | NO |
| **Thyroid dysfunction** | NO | NO | NO | NO | NO | YES |
| **SARS-CoV-2 swab** | Positive at admission | Positive at admission | Positive at admission | Positive at admission | Positive at second test | Positive at admission |
| **Pulmonary outcome** | Bilateral interstitial pneumonia due to COVID-19 | Bilateral interstitial pneumonia due to COVID-19 | Bilateral interstitial pneumonia due to COVID-19 | Bilateral interstitial pneumonia due to COVID-19 | Bilateral interstitial pneumonia due to COVID-19 and *E. Cloacae* | Bilateral interstitial pneumonia due to COVID-19 |
| **Hospitalization lenght (days)** | 16 | 29 | 27 | 38 | 18 | 7 |
| **Oxygen saturation (%)** | 79.7 | 92.2 | 92.2 | 94.1 | 87.0 | 90.5 |
| **Type of ventilation** | NIV/CPAP | CPAP/MV | NIV/MV | NIV/MV | NIV/CPAP/MV | NIV/CPAP/MV |
| **Pronation cycles** | Multiple | Multiple | Multiple | Multiple | Multiple | 2 |

Legend: LV = left ventricle; EF = ejection fraction; RCP = reactive c-protein; NIV = non-invasive ventilation; CPAP = continuous positive airway pressure; MV = mechanical ventilation.

### mRNA extraction and Digital PCR assay

Tissue samples for RNA were stored in RNAlater (Sigma-Aldrich, Saint Louis, MO, USA). Then, RNA was extracted from tissues by using TRIzol (ThermoFischer Scientific, Waltham, MA, USA) and Hybrid-R columns (GeneAll Biotechnology, Seoul, Korea), following the manufacturer’s instructions.

The RNA quantification was performed by spectrophotometer ND-1000 (NanoDrop®, EuroClone, Pero, Milan, Italy). For viral RNA quantification in myocardial specimens, a one-step chip-based digital PCR was performed on a QuantStudio 3D Digital PCR System platform composed by the QuantStudio 3D Instrument, the Dual Flat Block GeneAmp PCR System 9700 and the QuantStudio 3D Digital PCR Chip Loader (ThermoFischer Scientific, Waltham, MA, USA). TaqMan Fast Virus 1-Step Master Mix (Applied Biosystems, Foster City, CA, USA) was used in combination with TaqMan primers (10006830, 10006831) and FAM probe (10006832) for viral N gene purchased from Integrated DNA Technologies (Integrated DNA Technologies, Coralville, IA, USA), the primers and probe for the N gene were controlled and authorized by the Centers for Disease Control and Prevention, and human RPLP0 gene (Hs99999902_m1) with a VIC probe, as internal amplification control, purchased from Thermo Fisher Scientific. Reverse transcription (RT) was performed at 50°C for 10 minutes, followed by RT inactivation/initial denaturation at 96°C for 5 minutes.

PCR cycling consisted of a denaturation step at 98°C for 30 seconds followed by annealing/extension at 56°C for 1 minute (40 cycles) and a final extension at 60°C for 5 minutes. Analyses were executed with the online version of the QuantStudio 3D AnalysisSuite (ThermoFischer Scientific Cloud, Waltham, MA, USA) following the manufacturer’s instructions.

### Western blot analysis

Heart tissues were fragmented by Spectrum™ Bessman Tissue Pulverizers (Fisher Scientific, Hampton, NH, USA) and lysed in cell lysis buffer (Cell Signaling Technology, Danvers, MA, USA) supplemented with protease and phosphatase inhibitor cocktails (Sigma-Aldrich, Saint Louis, MO, USA).

Total protein extracts were quantified by BCA Protein Assay (ThermoFischer Scientific, Waltham, MA, USA), then subjected to SDS-PAGE and transferred onto a nitrocellulose membrane. The membranes were blocked for 1 hour at room temperature (RT) in Wash Buffer (Tris Buffer Sulfate 1×, 0.1% Tween 20) supplemented with 5% Bovine Serum Albumine (BSA) and then incubated over-night (O/N) at 4°C with the appropriate primary antibody. Primary antibodies adopted for western blot analysis were specific for ACE2 (R&D System, Minneapolis, MN, USA), SARS-CoV-2 nucleoprotein (Sino Biological, Düsseldorfer, Germany), SARS-CoV-2 spike protein (GeneTex, Irvine, CA, USA), and GAPDH (Santa Cruz Biotechnology, Dallas, TX, USA).

The membranes were incubated with peroxidase-conjugated secondary antibodies (GE Healthcare, Chicago, IL, USA) for 1 hour. Signals were visualized using the Western blotting chemiluminescence substrate LiteUP (EuroClone, Pero, Milan, Italy). Images were acquired with the ChemiDoc system (Bio-Rad Laboratories, Hercules, CA, USA).

### Immunostainings and histochemistry

Hearts samples were mapped as follows: full thickness samples from ventricles, atri (2 for each side), and 2 from interventricular septum. Specimens were routinely fixed in 10% buffered formaldehyde for 48 hours as recommended^23^ and then 3-μm sections were processed for histopathologic evaluation as well as for immunostainings.

Immunohistochemistry was performed with the automatic immunostainer DAKO OMNIS (DAKO, Glostrup, Denmark), using staining kit with magenta and brown chromogens, routinely applied in our laboratory. The magenta staining was adopted to avoid misinterpretation with lipofuscin granularity typical of cardiomyocyte cytoplasm.

The histochemistry with phosphotungstic acid haematoxylin (PTAH) was performed according to manufacturer’s protocol (BioOptica, Milan, Italy).

Primary antibodies adopted for immunohistochemistry and immunofluorescence were specific for SARS-CoV-2 nucleoprotein (Sino Biological, Düsseldorfer, Germany), SARS-CoV-2 spike protein (GeneTex, Irvine, CA, USA), and α-SARC (AbCam, Cambridge, UK).

As for SARS-CoV-2 nucleoprotein, samples from lungs of patients under study, with immunoreactivity in reactive pneumocytes, macrophages and epithelial respiratory cells were used as controls. Negative controls were also included in the study by omitting the primary antibody. For all the stainings, archival heart and lung tissue samples from patients dead for diseases other than COVID-19 and cardiologic causes were used as controls (Healthy Control).

Immunofluorescence images and analyses were performed by using a Zeiss LSM710 confocal microscope (Carl Zeiss, Oberkochen, Germany).

Except for immunofluorescence, all slides were digitalized with the NanoZoomer S360 Digital slide scanner C13220-01 (Hamamatsu Photonics, Milan, Italy).

### RNAscope assays

The RNAScope assays (ACD, Bio-technè, Minneapolis, MN, USA) was performed on 3-μm formalin-fixed paraffin-embedded sections, following Wang *et al*. published method^24^, by using specific probes for sense and antisense SARS-CoV-2 RNA spike sequences. Nuclei were counterstained with haematoxylin and the images were acquired by AxioImager microscope (Carl Zeiss, Oberkochen, Germany).

### TUNEL assay

DNA fragmentation associated to necrosis and/or apoptosis was visualized using a terminal deoxyribonucleotidyl transferase-mediated dUTP-digoxigenin nick end labeling (TUNEL) detection kit (Roche, Mannheim, Germany). At last, nuclei were counterstained with Hoechst (ThermoFischer Scientific, Waltham, MA, USA) and images were acquired by ApoTomell microscope (Carl Zeiss, Oberkochen, Germany).

### Transmission Electron Microscopy

Cardiac tissue samples for ultrastructural analysis were fixed in 2.5% glutaraldehyde in 0.13 M phosphate buffer pH 7.2-7.4 for 2 hours, post-fixed in 1% osmium tetroxide, dehydrated through graded ethanol and propylen oxide, and embedded in epoxy resin. Ultrathin sections of 50 to 60 nm not counterstained were observed with transmission electron microscope Leo 912ab (Carl Zeiss, Oberkochen, Germany).

### Ethical approval

Approval was not required (GU n. 72 26/03/2012, page 49).

## Results

### Patients’ clinical and autoptic features

The patient features reported in **Table 1** highlight that all the subjects died for respiratory failure. **Supplementary Figures 1** shows a representative result of a pulmonary CT, pathognomonic for COVID-19 disease. Only 1 out of 6 presented a slightly altered cardiac clinical profile, due to a mild hypertrophic LV, likely secondary to hypertension, occurring before the hospitalization for COVID-19 disease. Importantly, all patients did not show alterations of ultrasensitive Troponin I; 2 out of 6 were hypertensive; 2 out of 6 showed hypercholesterolemia, none was diabetic, and 1 presented thyroid dysfunction. All the 6 patients were assisted by mechanical ventilation in Intensive Care Unit for a period ranging from 6 to 21 days.

Concerning autopsy details, hearts ranged in size 410-750 grams (normal: 365 ± 71 grams), with cardiomegaly related to right ventricle dilation, due to lung overload. On cut surface, cardiac tissue was firm, brown, free of lesions, with some delicate areas of myocardiosclerosis in 1 out of 6 patient left ventricles; the coronary system was unremarkable, with no significant stenosis nor thrombosis.

### SARS-CoV-2 genome and proteins are present in cardiac samples of COVID-19 patients

We initially confirmed SARS-CoV-2 cardiac tropism in these patients, by examining with chip-based digital PCR the presence of viral RNA in total RNA extracts from heart tissue samples. Viral RNA was found in all COVID-19 heart specimens, but not in the healthy control. The number of copies of SARS-CoV-2 RNA molecules ranged from 4.44 to 5.33 log10(copies/mL) (**Figure 1a**).

**Figure 1.**
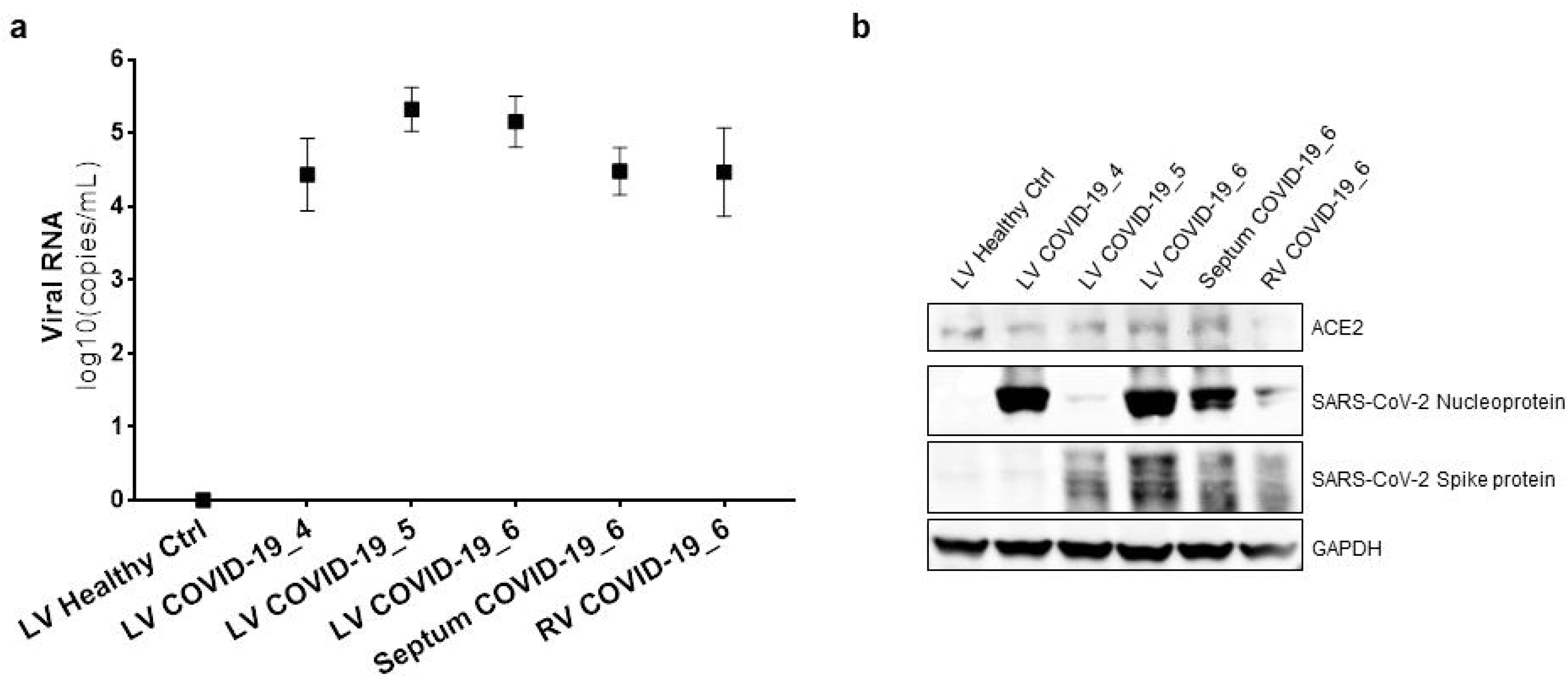
RNA and protein of SARS-CoV-2 are present in cardiac samples of COVID-19 patients. **a)** Chip-based digital PCR for SARS-CoV-2 N sequence in total RNA extracts from tissues of 3 different patients and different cardiac regions. Raw expression values were expressed as log10(copies/mL). The results are expressed as median and confidence interval (CI 95%). **b)** Western blot analysis for ACE2, SARS-CoV-2 nucleoprotein, and spike protein on total protein extracts from tissues of 3 put of 6 different patients and different cardiac regions. GAPDH has been adopted as loading control. LV, left ventricle; Septum, interventricular septum; RV, right ventricle.

A Western blot analysis of total protein cardiac autopsy specimen lysates confirmed that the well-known viral gate of host cells ACE2 receptor^17^ was expressed in the cardiac tissue^20^. Similar levels of expression were found in different cardiac districts and, of note, independently on COVID-19. Furthermore, we showed, for the first time, that SARS-CoV-2 viral proteins are detectable in COVID-19 patient hearts, while no traces have been found in the healthy control. In detail, nucleoprotein and spike protein were expressed at different levels in the analysed patients (**Figure 1b**), raising the hypothesis that the viral genome is actively transcribed into cardiomyocytes.

### SARS-CoV-2 is localized in cardiomyocytes of COVID-19 patients in terms of viral proteins and RNA

The light microscopy analyses performed firstly allowed us to define all COVID-19 cases as similar by microscopic aspects. In details, the immunohistochemical analyses confirmed positivity for COVID-19 in left ventricular cardiac samples for SARS-CoV-2 nucleoprotein and spike protein (**Figure 2a, b**). Of note, viral proteins were found within cardiomyocytes cell body, and more specifically, in cytosolic are of lipofuscin (**Suppl. Fig. 2a**). This finding was confirmed by immunofluorescence co-staining of viral nucleoprotein with αSARC-positive cells (**Figure 2c**).

**Figure 2.**
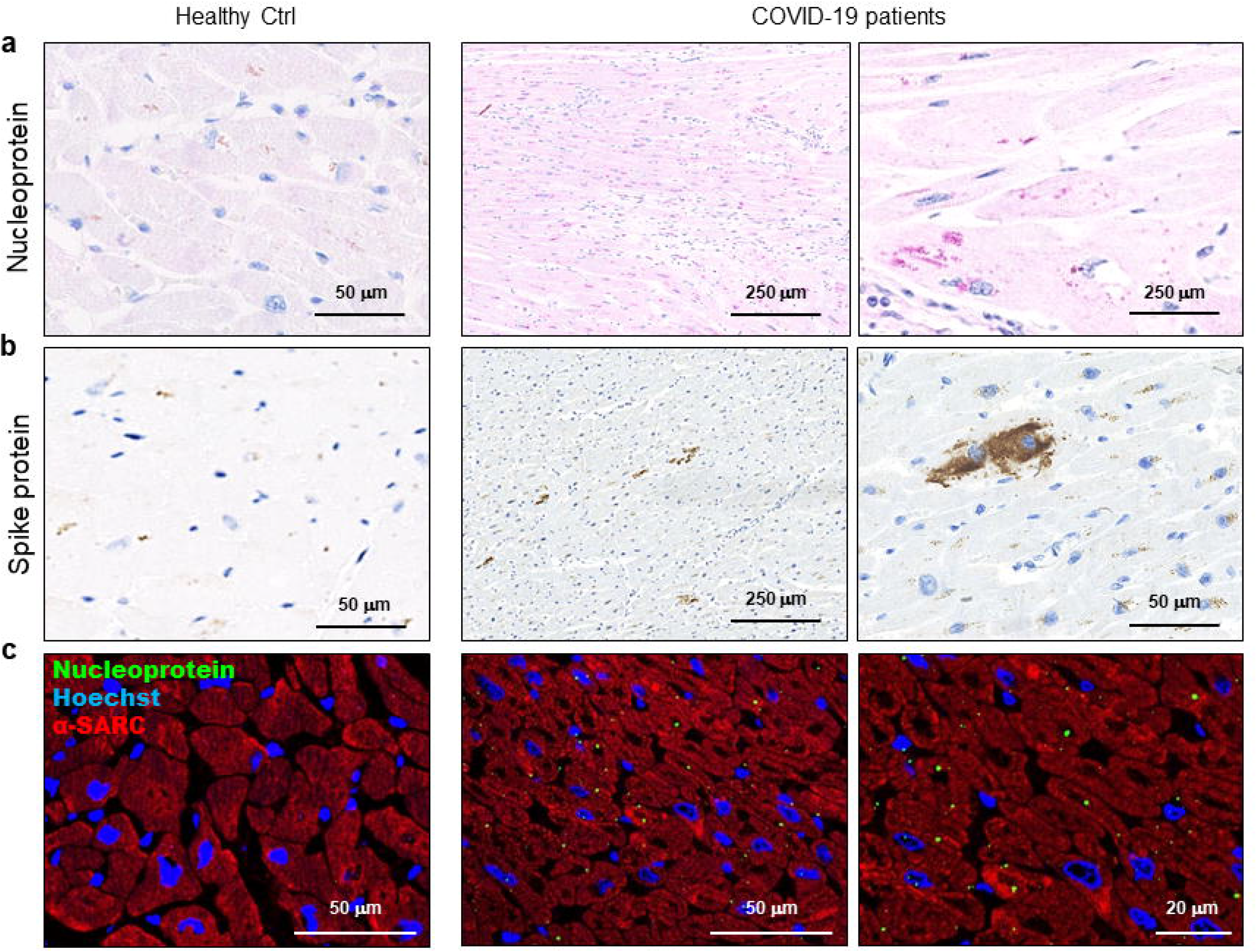
SARS-CoV-2 proteins are detectable in cardiomyocytes of COVID-19 patients. **a-b)** Representative images of immunohistochemistry assays on 3μm-slides of formalin-fixed paraffin-embedded left ventricle specimens from healthy control (Healthy Ctrl, left panels) and COVID-19 patients (COVID-19, central and right panels) for SARS-CoV-2 nucleoprotein (**a**, in red) and spike protein (**b**, in brown). **c)** Representative images of immunofluorescence assay on 3μm-slides of formalin-fixed paraffin-embedded left ventricle specimens from healthy control (Healthy Ctrl, left panels) and COVID-19 patients (COVID-19, central and right panels) for SARS-CoV-2 nucleoprotein (green) and sarcomeric α-actin (α-SARC, red). Nuclei have been stained by Hoechst (blue). Magnification as scale bars.

In order to determine the presence of SARS-CoV-2 as transcriptionally active virus into cardiomyocytes, we performed the RNAScope assay (**Figure 3**).^24^ In detail, this *in situ* hybridization was performed by use of two different probes, recognising the spike protein RNA, sense (**Figure 3a**) and antisense (**Figure 3b**), allowing distinction of the viral RNA genome from its transcript. Signals were obtained with both probes in cardiomyocytes of COVID-19 patients only, but the most abundant staining resulted from the probe detecting the actively transcribed virus (**Figure 3b**).

**Figure 3.**
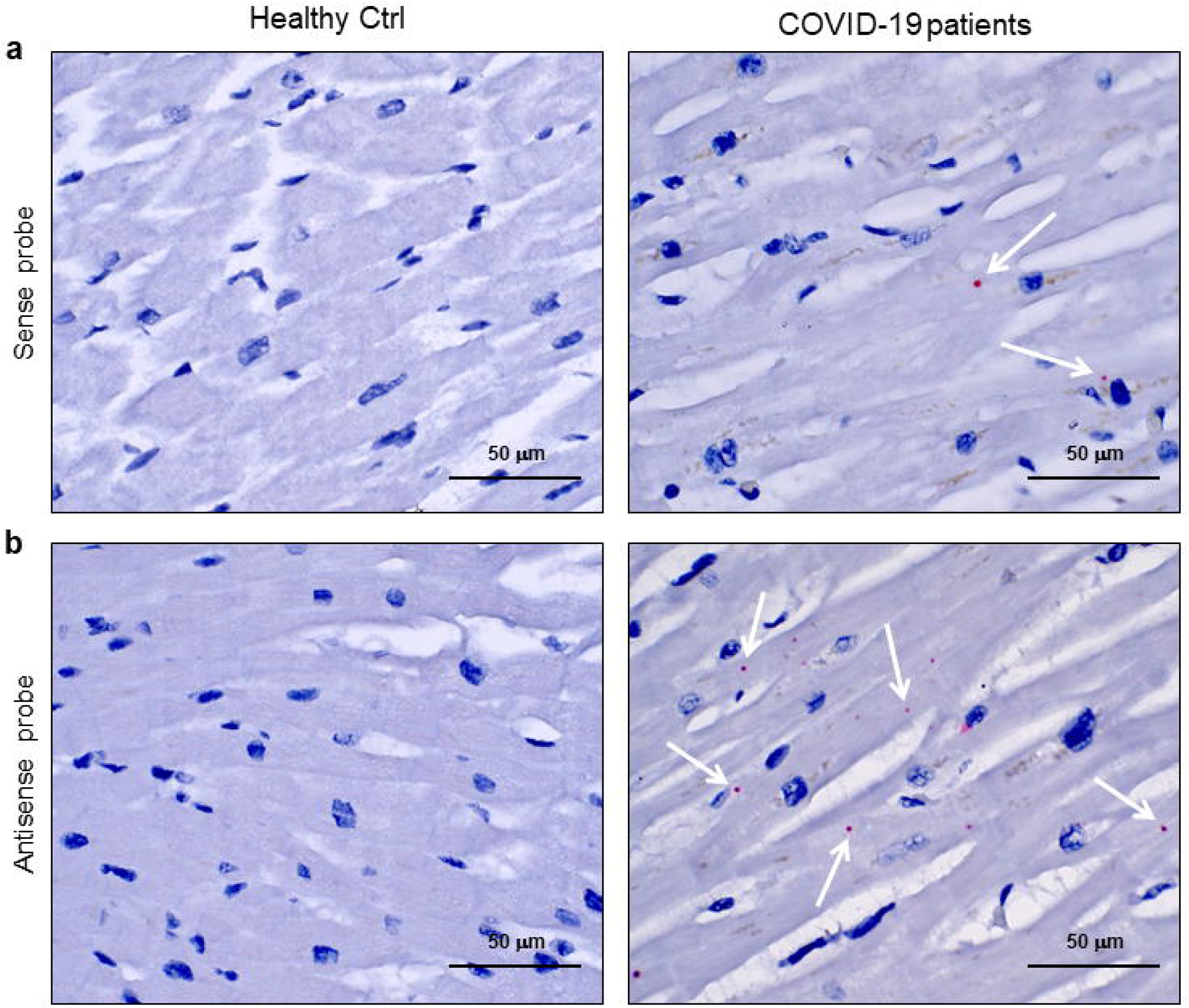
SARS-CoV-2 sense and antisense RNA are localized into cardiomyocytes of COVID-19 patients. **a-b)** Representative images of spatially resolved viral RNA detection by RNAScope assay on 3μm- slides of formalin-fixed paraffin-embedded left ventricle specimens from healthy control (Healthy Ctrl, left panels) and COVID-19 patients (COVID-19, central and right panels) for SARS-CoV-2 sense (**a**) and antisense (**b**) probes for spike protein RNA sequences. Fast Red dots indicate viral RNA presence (white arrows). Nuclei have been counterstained with haematoxylin. Magnification as scale bars.

### Cardiomyocytes containing SARS-CoV-2 do not show cell death sign, but display intracellular alterations

In **Figure 4**, the immunohistochemistry for SARS-CoV-2 nucleoprotein and PTAH staining, performed on consecutive heart sections, highlighted several microscopic scenarios. Not all the cardiomyocytes contain the virus, and among those positive for the viral nucleoprotein we found living and unaltered cells (**Figure 4a**). Nonetheless, we appreciated, through PTAH staining, isolated or grouped cardiomyocytes showing atypical degenerative aspects, such as increased cell volumes with intracellular oedema, together with rarefied, disarranged, disrupted, and plurifocally clumped myofibrils (**Figure 4b**).

**Figure 4.**
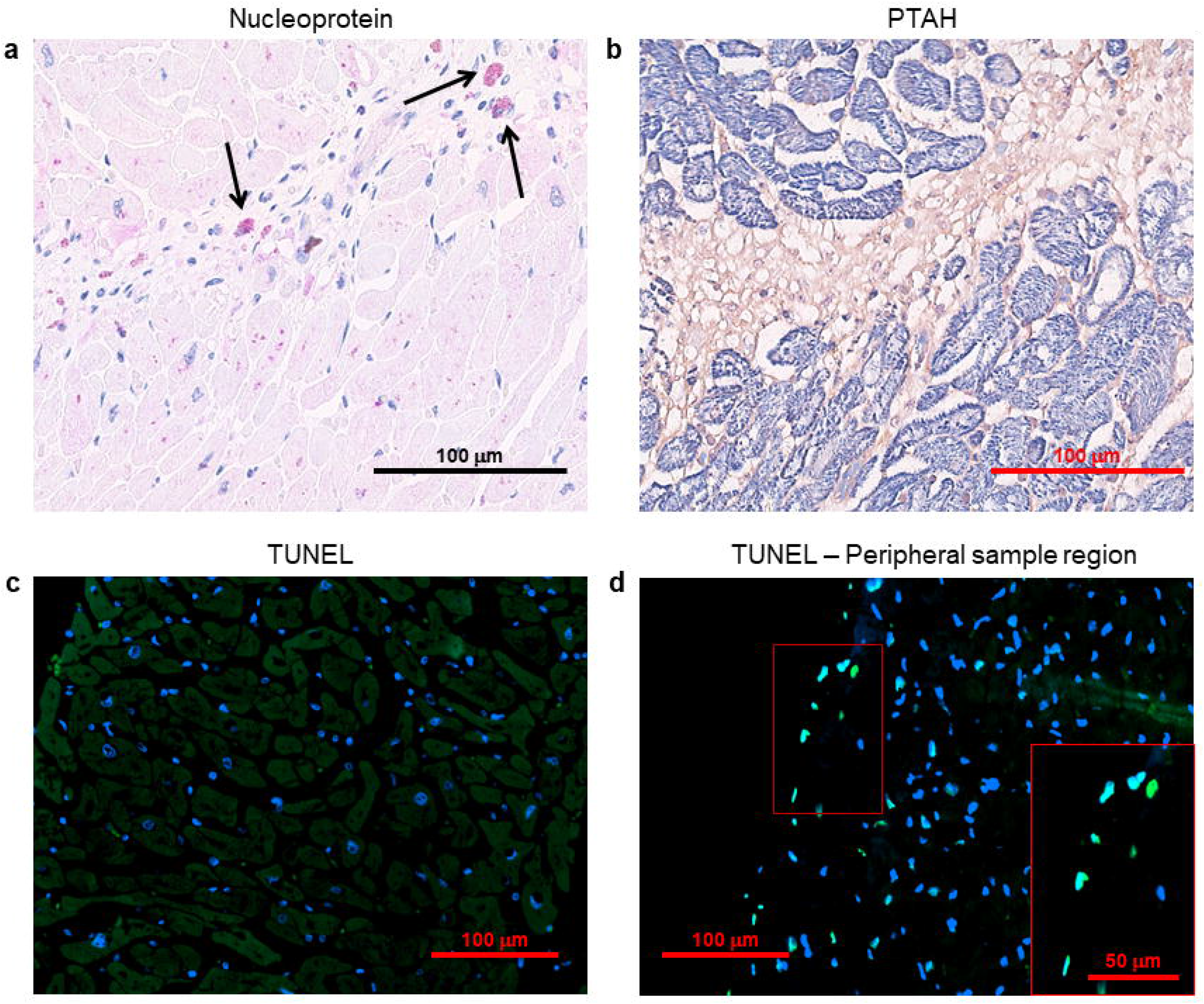
Several cardiomyocytes containing SARS-CoV-2 nucleoprotein display intracellular alterations, but not cell death signs. **a-b)** Representative images on consecutive slides of immunohistochemistry assay for SARS-CoV-2 nucleoprotein (**a**) and PTAH staining (**b**, sarcomere structures in violet/blue), performed on left ventricle specimens from COVID-19 patients. Black arrows indicate strongly immunoreactive interstitial cells for SARS-CoV-2 nucleoprotein (in red), with a morphology consistent with macrophages. **c)** Representative images of TUNEL assay on 3μm-slides of formalin-fixed paraffin-embedded left ventricle specimens from COVID-19 patients. Damaged DNA is marked in green, nuclei have been counterstained with Hoechst (blue). Magnification as scale bars.

In order to investigate whether these interesting features, concerning cardiac cell alterations, may be linked to apoptotic/necrotic processes activation, we performed TUNEL assay on all the subject samples and we detected no cell death signs, except for the peripheral sample regions (**Figure 4c, d**).

Vascular modifications were unremarkable; no endothelial viral cytopathic effects nor endotheliitis was here documented. Furthermore, a low grade of inflammatory infiltrates was observed in cardiac tissue, especially in sub-pericardiac regions without a specific distribution (**Suppl. Fig. 2b**). Sporadic interstitial macrophages displayed a strong immunoreactivity for SARS-CoV-2 nucleoprotein (**Suppl. Fig. 2c**).

The investigation by electron microscopy confirmed the previously obtained evidence on the SARS-CoV-2 presence in cardiomyocytes. Indeed, single and/or grouped spherical particles about ~130 nm in diameter, with electron dense periphery characterized by crown-shaped small external projections and a morphology compatible to SARS-CoV-2 were clearly visible in the intracellular space between sarcomeres and sarcolemma of cardiomyocytes (arrows, **Figure 5a, b**).

**Figure 5.**
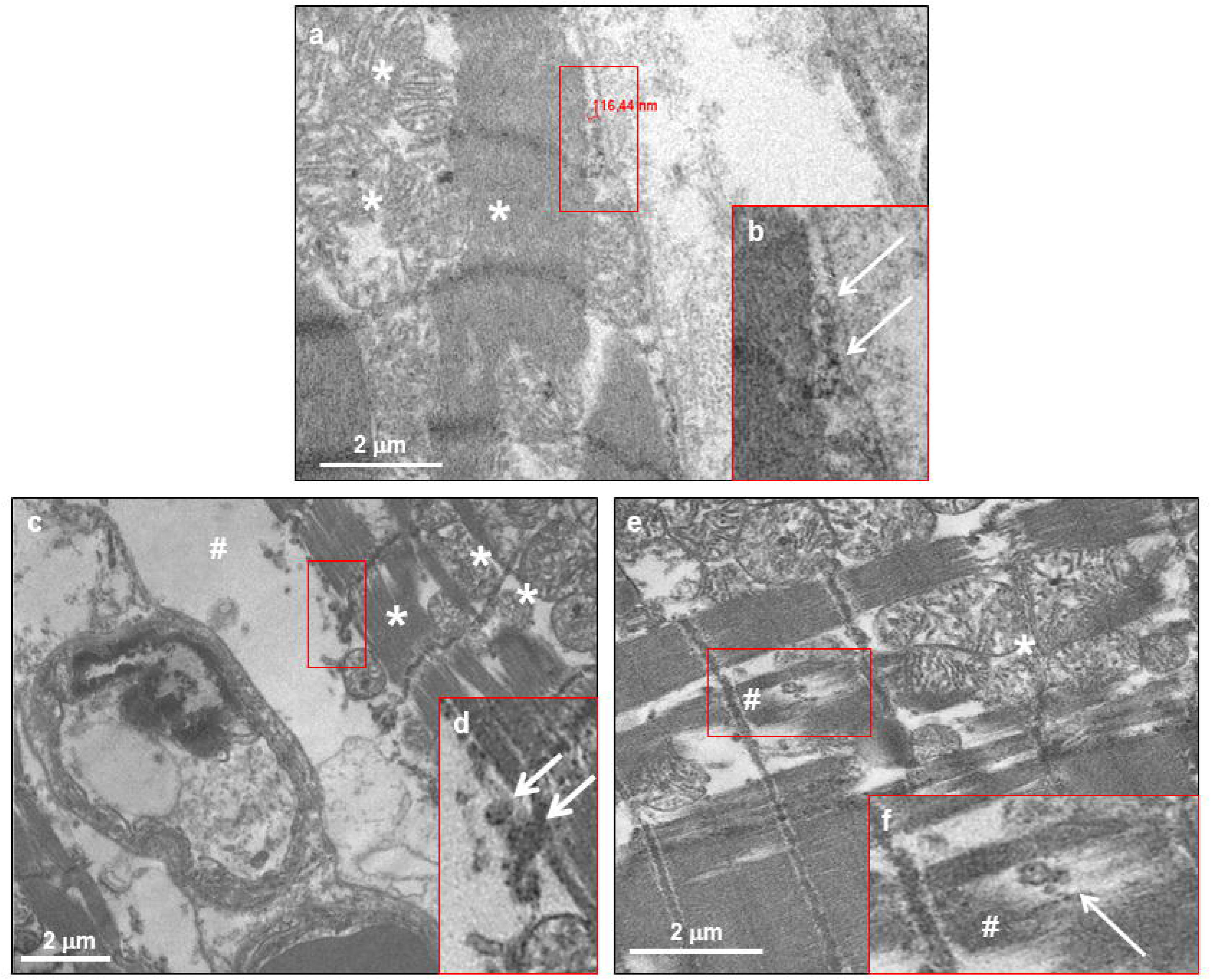
SARS-CoV-2 virions are localized within both normal and altered cardiomyocytes of COVID-19 patients. **a, c, e)** Electron micrograph showing cardiomyocytes containing viral particles (white arrows), compatible with crown-shaped SARS-CoV-2. The viral particle diameter (**a**) is of ~116 nm. **b, d, f)** High magnifications of red squared areas of panel a, c, and e, respectively. In all panels, asterisks indicate normal mitochondrial and sarcomere structures, while cytoplasmic and sarcomere alterations are indicated with hashtags.

As for immunohistochemistry and histology assays, the cardiomyocyte phenotype is not univocal: while some cardiomyocyte hosting SARS-CoV-2 did not present any evident abnormality in terms of cellular structures, other cardiac cells presented peculiar degenerative features, such as excessive intracellular area between sarcolemma and sarcomere structure (**Figure 5c, d**) and abnormal, disrupted and clumped myofibrils (**Figure 5e, f**). Regarding the former alteration, this cardiomyocyte swelling (**Figure 5c**, hashtag) may microscopically explain the cardiac oedema, previously observed in histological examinations. Noteworthy, the sarcomere rupture occurs in co-localization of SARS-CoV-2 viral particle (**Figure 5f**, hashtag).

Although the microscopy investigation was compatible with cardiomyocyte damage, it is important to highlight that, at ultrastructural level, the sarcolemma and mitochondria resulted unaltered and structurally undamaged (**Figure 5a, c, e**, asterisks), thus not suggestive of cardiomyocyte death.

## Discussion

In this work, we reported the presence of SARS-CoV-2 in the cardiac tissue of a series of 6 autoptic samples, obtained from patients died for respiratory failure with no clinical signs of cardiac involvement and with no heart disease history. The originality of our study consists in the observation, for the first time, in cardiomyocytes of i) SARS-CoV-2 RNA and protein localization and ii) active viral transcription of both sense and antisense RNA for SARS-CoV-2 spike protein.

Although several papers have already reported the presence of SARS-CoV-2 RNA in the heart,^12,15,16^ none of them has detected viral localization in cardiomyocytes. We here demonstrated, by means of several techniques, the undisputable presence of active SARS-CoV-2 in COVID-19 patients’ cardiomyocytes, in terms of viral transcript detection (by digital PCR and RNAScope assays), viral protein finding (by Western blot, immunohistochemistry and immunofluorescence), and full virions identification (by electron microscopy). Specifically, a clear signal for the SARS-CoV-2 spike protein was found in cardiomyocytes only, while a granular diffuse immunostaining of the SARS-CoV-2 nucleoprotein antigen was present both in cardiomyocytes and in interstitial inflammatory cells, mostly in macrophages. This latter data is in line with viral particles detection in cardiac CD68^+^ cells described by Tavazzi *et al*.^11^ and with RNA viral presence in macrophages recently reported by Lindner and colleagues,^12^ but added unprecedented insights about SARS-CoV-2 protein expression and localization in the heart.

In this work, we have also observed intracellular alterations in cardiomyocytes harbouring SARS-CoV-2. To date, studies based on endomyocardial biopsies or isolated autopsy description did not reveal cardiomyocyte alterations in presence of SARS-CoV-2.^11-13,20^ In our samples, scattered and focal degenerative cardiomyocyte features were confirmed by electron microscopy image analyses at ultrastructural level. In details, several cardiomyocytes showed an altered cell structure, with increased cell area and intracellular oedema, suggesting a cardiomyocyte swelling. Moreover, damaged sarcomeres, with disrupted and clumped myofilaments, were also observed, in some cases associated to rarefaction of myofibrils. It is important to point out that none of these cells showed disrupted or damaged sarcolemma. As for cardiac cell swelling, this microscopic event may be interpreted as a precocious subclinical sign of cell damage, potentially leading to myocardial interstitial oedema formation.^25^ Moreover, focal myofibrillar lysis are in line with observation previously described by Tavazzi *et al*.^11^ and might correspond to the atypical degenerative characteristics reported in myocytes from COVID-19 patient autoptic tissues.^26^ The small number of virions we observed is in line with the low SARS-CoV-2 loads, previously found by Escher *et al*.^14^ in a series of endomyocardial biopsies from patients with active heart disease, and with the reported absence of immunoreactivity for SARS-CoV nucleocapsid protein, described by Adachi *et al*. in a cardiac autopsy case report.^22^

Noteworthy, available histologic findings reported in literature often lack a correlation to clinical data *(e.g*. cardiac troponin levels), thus clinical relevance and incidence of COVID-19 cardiac involvement still remains unclear. Our data complete the current literature in suggesting that the cardiac tissue, including the contractile cell compartment, can be infected by the SARS-CoV-2 virus, and this may initially result in microscopic cardiomyocyte alterations, not leading to clinically-relevant macroscopic cardiac damage.

Our crucial study specifically suggest the need to redefine post-discharge surveillance guidelines for surviving COVID-19 patients’ follow-up even in the absence of overt cardiac phenotype, since the cardiovascular long-term outcome of COVID-19 patients may mirror SARS patients, who developed cardiovascular abnormalities in 40% of recovered cases in a 12-years-long follow-up period.^5^

## Limitation of the study

This study has limitations inherent to the limited number of patients’ autopsies included. However, our findings clearly and consistently show that SARS-CoV-2 is present and active in cardiomyocytes of our COVID-19 patients. Accordingly, we deliberately did show any statistical analysis, since we observed a complete absence of SARS-CoV-2 signal in healthy control samples included in the experiments. For all these reason, future studies will be necessary to confirm our data which are, at present, unprecedented.

## Data Availability

The raw dataset is deposited in Zenodo web site, and available after e-mail request to the corresponding author.

https://dx.doi.org/10.5281/zenodo.3974891

## Acknowledgements

Transmission electron microscopy images were acquired by NOLIMITS, an advanced imaging facility of University of Milan.

## Author contribution

GB, GLP, SC and GP conceived of the work. GB, GLP, MF, DT, CM, PS performed the experiments and analysed the data. MF, ES, PP, SC, GP contributed with data interpretation. GLP, MF, ES drafted the manuscript, GP substantively revised it. All authors revised the manuscript, approved its final version and agreed to be accountable for all aspects of the work.

